# Development and validation of the pharmacological statin-associated muscle symptoms risk stratification (PSAMS-RS) score using real-world electronic health record data

**DOI:** 10.1101/2023.08.10.23293939

**Authors:** Boguang Sun, Pui Ying Yew, Chih-Lin Chi, Meijia Song, Matt Loth, Yue Liang, Rui Zhang, Robert J. Straka

**Author notes:** **Addresses for Correspondence:** Robert J. Straka, Pharm.D., Experimental and Clinical Pharmacology, University of Minnesota College of Pharmacy, 7-115B Weaver-Densford Hall, Minneapolis, Minnesota 55455 USA. **Author Contributions:** BGS conceived and designed the study, conducted data analysis, interpreted the results, and drafted and revised the paper. PYY, CLC, MJS, YL, RZ contributed to study design, data acquisition, results interpretation, and paper revision. RJS, CLC, RZ reviewed and further contributed to the manuscript. All authors drafted and revised the manuscript.

## Abstract

**Introduction:** Statin-associated muscle symptoms (SAMS) contribute to the nonadherence to statin therapy. In a previous study, we successfully developed a pharmacological SAMS (PSAMS) phenotyping algorithm that distinguishes objective versus nocebo SAMS using structured and unstructured electronic health records (EHRs) data. Our aim in this paper was to develop a pharmacological SAMS risk stratification (PSAMS-RS) score using these same EHR data.

**Method:** Using our PSAMS phenotyping algorithm, SAMS cases and controls were identified using University of Minnesota (UMN) Fairview EHR data. The statin user cohort was temporally divided into derivation (1/1/2010 to 12/31/2018) and validation (1/1/2019 to 12/31/2020) cohorts. First, from a feature set of 38 variables, a Least Absolute Shrinkage and Selection Operator (LASSO) regression model was fitted to identify important features for PSAMS cases and their coefficients. A PSAMS-RS score was calculated by multiplying these coefficients by 100 and then adding together for individual integer scores. The clinical utility of PSAMS-RS in stratifying PSAMS risk was assessed by comparing the hazard ratio (HR) between 4th vs 1st score quartile.

**Results:** PSAMS cases were identified in 1.9% (310/16128) of the derivation and 1.5% (64/4182) of the validation cohort. After fitting LASSO regression, 16 out of 38 clinical features were determined to be significant predictors for PSAMS risk. These factors are male gender, chronic pulmonary disease, neurological disease, tobacco use, renal disease, alcohol use, ACE inhibitors, polypharmacy, cerebrovascular disease, hypothyroidism, lymphoma, peripheral vascular disease, coronary artery disease and concurrent uses of fibrates, beta blockers or ezetimibe. After adjusting for statin intensity, patients in the PSAMS score 4th quartile had an over seven-fold (derivation) (HR, 7.1; 95% CI, 4.03-12.45) and six-fold (validation) (HR, 6.1; 95% CI, 2.15-17.45) higher hazard of developing PSAMS versus those in 1st score quartile.

**Conclusion:** The PSAMS-RS score can be a simple tool to stratify patients’ risk of developing PSAMS after statin initiation which can facilitate clinician-guided preemptive measures that may prevent potential PSAMS-related statin non-adherence.

## Introduction

Approximately 50% of individuals aged 65 and above in the United States use statins for prevention and/or treatment of cardiovascular disease.^1^ However, a significant proportion of statin users, ranging from 25% to 50%, do not fully experience statin benefits of reducing morbidity and mortality due to discontinuation.^2^ Various factors contribute to statin discontinuation with around 25% of former statin users citing side effects, particularly statin-associated muscle symptoms (SAMS), as the main reason for their non-adherence or discontinuation.^2,3^

SAMS signals are routinely documented in electronic health record (EHR) in structured formats including ICD codes for muscle symptoms, statin allergies, creatine kinase (CK) elevation, and also in unstructured formats such as a clinicians’ narrative description of SAMS found in clinical notes. Clinical phenotyping algorithms to identify SAMS have been developed in diverse EHR systems using structured data alone but also in some cases, both structured and unstructured data.^4–6^

However, the prevalence estimates of pharmacological SAMS can be confounded by the nocebo effect.^7^ As a result, the reported prevalence of SAMS can range from <1% in clinical trials to up to 25% from observational studies.^3^ Previously, our research team accomplished the development of a PSAMS phenotyping algorithm based on the University of Minnesota (UMN) Fairview EHR system.^8^ Our PSAMS algorithm, developed based on an annotated gold-standard set using the SAMS-Clinical Index (SAMS-CI) tool^9^, is capable of specifically phenotyping non-nocebo PSAMS.^8^ This approach allowed for a more precise identification of objective muscle symptoms pharmacologically related to statin use.

Patients’ demographics, genetics, concurrent drug use and comorbid conditions can contribute to PSAMS occurrences.^3^ For example, female gender is associated with a 3.1-fold increased risk of statin-induced myopathy and consequential discontinuation of statin therapy.^10,11^ However, these factors alone might have a low predictive value for PSAMS and thus there is a critical need for a multifactorial PSAMS risk stratification tool.^12,13^ So far, limited studies have been conducted to propose a prognostic risk stratification score integrating a variety of clinical factors associated with PSAMS. Therefore, in this study, we aimed to create a PSAMS risk stratification (PSAMS-RS) score that can identify patients with a significantly higher risk of developing PSAMS based on their clinical factors. The precise identification of PSAMS cases and controls was accomplished by our PSAMS phenotyping algorithm.^8^ The development of the PSAMS-RS score can enable healthcare providers to proactively identify individuals who may be at higher risk of PSAMS and thereby tailor their treatment approaches accordingly to improve statin adherence.

## Methods

### Data Sources and Processing

As described in our PSAMS phenotyping study, the statin user cohort was retrieved from the UMN Fairview EHR between 1/1/2010 to 12/31/2020. Briefly, the statin cohort contains patients over 18 years old at index date (the day the patient was prescribed their first statin), were regular UMN Fairview system users but not prevalent statin users. The follow-up period was 1 year after the index date or the end of the study period, whichever was earlier. The study is approved by University of Minnesota IRB (STUDY00011134).

### Potential Predictive Variables

Potential predictive variables included the following patient characteristics according to various literature^3,14–16^: demographic variables at index date, medical histories within one year prior to index date and concurrent medications within three months prior to index date (Table 1).

**Table 1:**
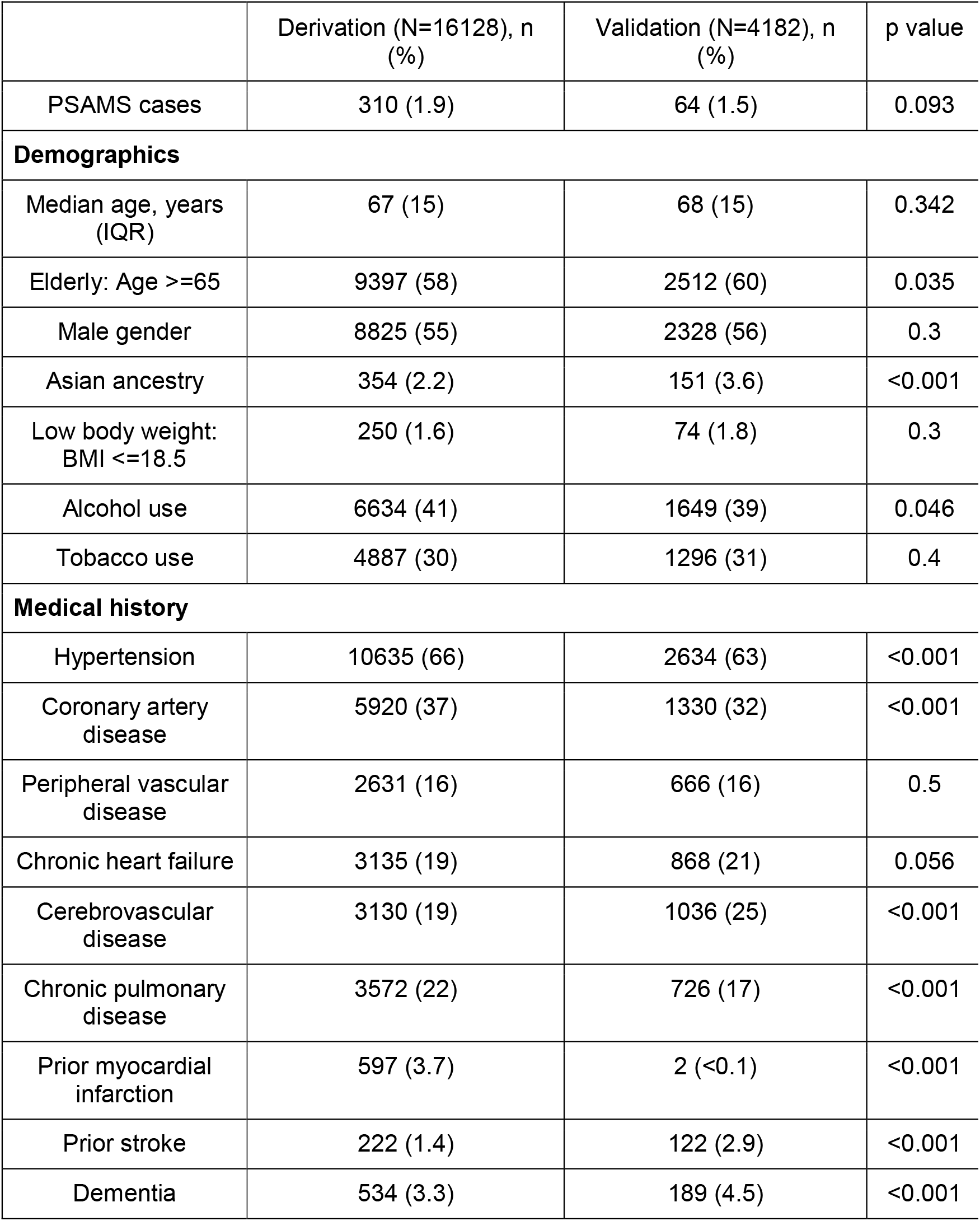
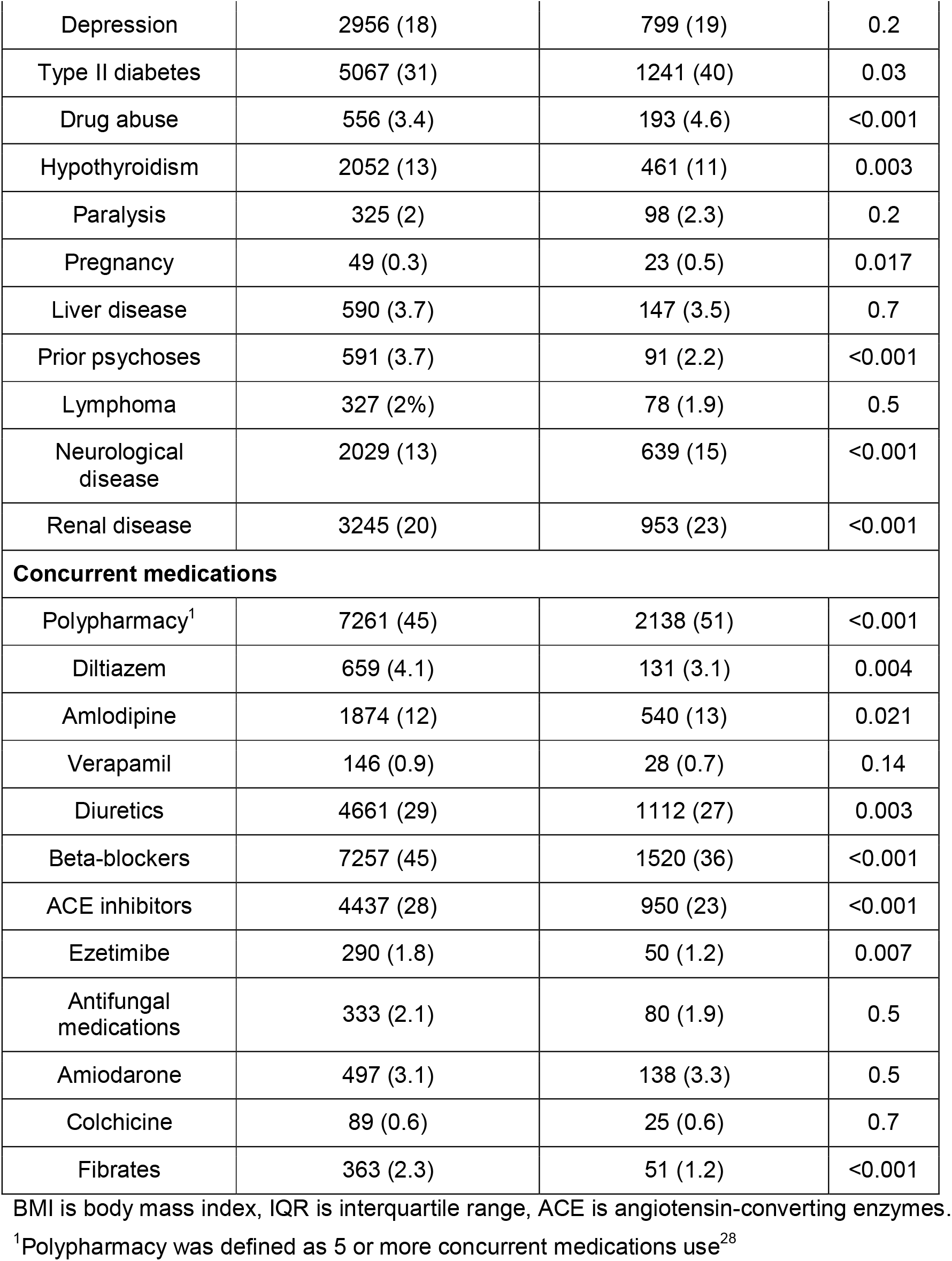
Baseline Characteristics in the Derivation and Validation Set.

### Outcomes

The primary outcome in the study was binary PSAMS at one year of statin initiation. The PSAMS phenotyping algorithm was utilized to identify PSAMS cases. Briefly, the PSAMS phenotyping algorithm is a rule-based algorithm that incorporates ICD codes, allergy list, CK elevation and clinical notes mentions of muscle pain associated with statins. The final combined rule-based algorithm considers both clinical notes and structured data criteria in a hierarchical manner with a precision of 0.85, recall of 0.71, and F1 score of 0.77 against the gold standard set created by expert annotations.

### Model Development

The statin cohort was temporally split into derivation (1/1/2010 to 12/31/2018) and validation (1/1/2019 to 12/31/2020) cohorts.^17^ The derivation cohort was further split into training (80%) and testing (20%) cohorts for model development. 38 variables were identified from EHR data and were used in the variable selection process using the Least Absolute Shrinkage and Selection Operator (LASSO) regression. The response variable being modeled is the binary PSAMS. The LASSO regression process has been shown to have superior predictive model selection performance than conventional logistic regressions.^18,19^ The hyperparameter λ (penalty) in the LASSO regression was chosen based on the best model discrimination ability per area under the receiver-operator characteristic curve (ROC-AUC) on the training set with 10-fold cross validation.^20^ A PSAMS-RS score is obtained by multiplying each β coefficient obtained from the LASSO regression (using the chosen λ) by 100 and rounding them to the nearest integer for easier clinical interpretation.^21^ The multiplying integer (100) was chosen based on its ability to transform the LASSO coefficients to integers without excessive roundings. These resulting integer values for all relevant variables were summed together to calculate a total score for each patient.^22^ An baseline integer score was added to prevent negative score values and potential complexities in clinical interpretation. Additionally, selective machine learning classifiers were fitted using the selected variables from LASSO regression to predict the binary PSAMS. These machine learning classifiers included Gaussian Naive Bayes, Support Vector Machines, Adaptive Boosting, Gradient Boosting and Random Forest. The hyperparameters for the machine learning algorithms were kept at default values because the performances of these models only served as references for the main interpretable LASSO model.

### Model Performance and Validation

The model performance of multifactorial predictors selected from LASSO regression was examined by fitting them into a logistic regression with the binary PSAMS as outcome.^23,24^ The model’s discrimination ability in the derivation and validation set were assessed using ROC-AUC scores, and the model calibration was assessed using Hosmer-Lemeshow goodness-of-fit test.^25^ The performance of the selective machine learning algorithms were evaluated on the testing set. Additionally, the performance of the Myopathy Risk Score (MRS) from Hopewell et al. was also assessed using the derivation cohort.^26^

The PSAMS-RS score was discretized into four equal quantiles (quartiles) to facilitate risk stratification. Furthermore, a PSAMS-RS score binary cutoff was determined to provide an even faster risk stratification strategy. The optimal binary score cutoff was determined by optimizing the balance of precision (the proportion of true positive predictions out of all positive predictions, measuring the accuracy of positive predictions for PSAMS cases) and recall (the proportion of true positive predictions out of all actual positive instances, measuring the ability to identify all true positive PSAMS cases). To assess the PSAMS-RS score’s ability to identify patients at high PSAMS risk within one year of statin initiation, Kaplan-Meier curves and Cox Proportional Hazard models were utilized. The one-year followup was chosen because it was the timeframe when most statin adverse events occur.^2^ The development and validation of this risk model followed the Transparent Reporting of a Multivariable Prediction Model for Individual Prognosis or Diagnosis (TRIPOD) statement.^27^

## Results

Table 1 presents the baseline demographic and clinical characteristics of the 20310 patients included in the study. The cohort identification flow diagram from our previous PSAMS phenotyping study provides a detailed overview of the patient selection process.^8^ Among the final included patients, 1.9% (310/16128) in the derivation and 1.5% (64/4182) in the validation cohort were identified as PSAMS cases.

Out of the 38 variables considered in the LASSO regression, 16 variables were found to be significant in predicting PSAMS after optimizing the λ (penalty) value. Table 2 displays the multivariate regression results for these 16 variables, along with their corresponding LASSO β coefficients and point scores. Table S1 displays the ICD definitions for comorbidity variables included in the score. The comorbidity variables were defined based on Charlson^29^ and Elixhauser Comorbidity^30^ Index and their ICD coding was derived from the Quan 2005 coding algorithm.^31^ A user-friendly PSAMS-RS score calculator was provided in the appendix.

**Table 2:**
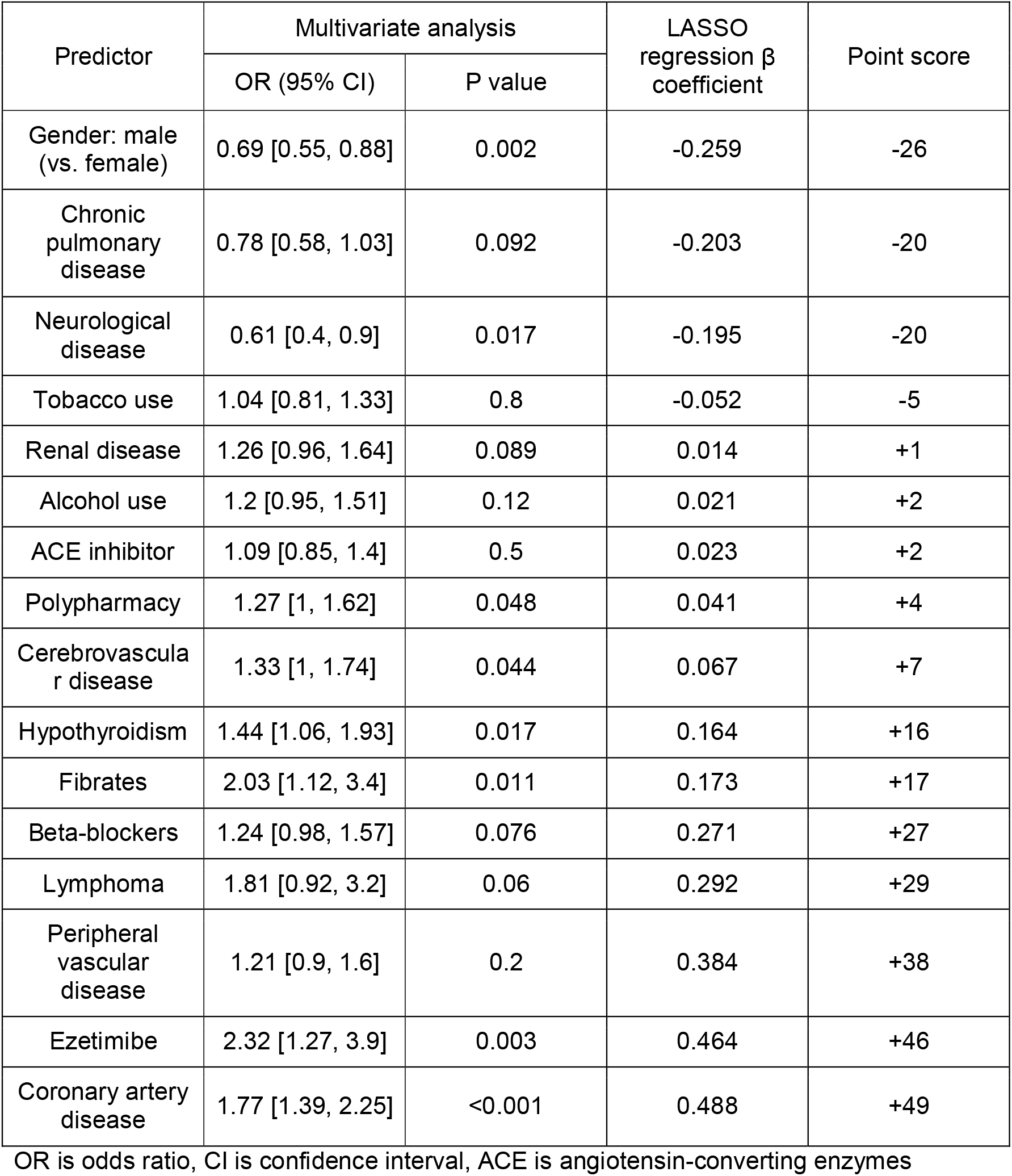
Pharmacological Statin-Associated Muscle Symptoms-Risk Stratification (PSAMS-RS) Score.

| Predictor | Multivariate analysis | | LASSO regression $\beta$ coefficient | Point score |
| --- | --- | --- | --- | --- |
|  | OR (95% CI) | P value |  |  |
| Gender: male (vs. female) | 0.69 [0.55, 0.88] | 0.002 | -0.259 | -26 |
| Chronic pulmonary disease | 0.78 [0.58, 1.03] | 0.092 | -0.203 | -20 |
| Neurological disease | 0.61 [0.4, 0.9] | 0.017 | -0.195 | -20 |
| Tobacco use | 1.04 [0.81, 1.33] | 0.8 | -0.052 | -5 |
| Renal disease | 1.26 [0.96, 1.64] | 0.089 | 0.014 | +1 |
| Alcohol use | 1.2 [0.95, 1.51] | 0.12 | 0.021 | +2 |
| ACE inhibitor | 1.09 [0.85, 1.4] | 0.5 | 0.023 | +2 |
| Polypharmacy | 1.27 [1, 1.62] | 0.048 | 0.041 | +4 |
| Cerebrovascular disease | 1.33 [1, 1.74] | 0.044 | 0.067 | +7 |
| Hypothyroidism | 1.44 [1.06, 1.93] | 0.017 | 0.164 | +16 |
| Fibrates | 2.03 [1.12, 3.4] | 0.011 | 0.173 | +17 |
| Beta-blockers | 1.24 [0.98, 1.57] | 0.076 | 0.271 | +27 |
| Lymphoma | 1.81 [0.92, 3.2] | 0.06 | 0.292 | +29 |
| Peripheral vascular disease | 1.21 [0.9, 1.6] | 0.2 | 0.384 | +38 |
| Ezetimibe | 2.32 [1.27, 3.9] | 0.003 | 0.464 | +46 |
| Coronary artery disease | 1.77 [1.39, 2.25] | <0.001 | 0.488 | +49 |

OR is odds ratio, CI is confidence interval, ACE is angiotensin-converting enzymes

**PSAMS-RS score** = 71 - 26 * male gender - 20 * chronic pulmonary disease - 20 * neurological disease - 5 * tobacco use + 1 * renal disease + 2 * alcohol use + 2 * ACE inhibitor use + 4 * polypharmacy + 7 * cerebrovascular disease + 16 * hypothyroidism + 17 * fibrates use + 27 * beta blockers use + 29 * lymphoma + 38 * peripheral vascular disease + 46 * ezetimibe + 49 * coronary artery disease

Figure 1 illustrates the observed rates of PSAMS at one year of statin initiation based on the PSAMS-RS score quartiles in both the derivation and validation cohorts. The PSAMS-RS score demonstrates a significant gradient in the distribution of PSAMS, with similar patterns observed in both derivation and validation cohorts (p for trend <0.05 for both). When the binary PSAMS score cutoff is 102, the precision and recall of differentiating PSAMS cases and controls were optimized. The proportions of PSAMS in patients with PSAMS-RS scores >=102 and <102 at one year were 3.1% (180/6055) versus 1.2% (130/10073) in the derivation cohort (p <0.001) and 2.2% (30/1354) versus 1.2% (34/2828) in the validation cohort (p =0.018), respectively. Overall, 37.5% (6055/16128) patients in the derivation cohort and 32.4% (1354/4182) in the validation cohort had PSAMS-RS scores >=102. Additionally, 2.6% (420/16128) patients in the derivation cohort and 2% (82/4182) in the validation cohort were at 4th quartile of the PSAMS-RS scores.

**Figure 1:**
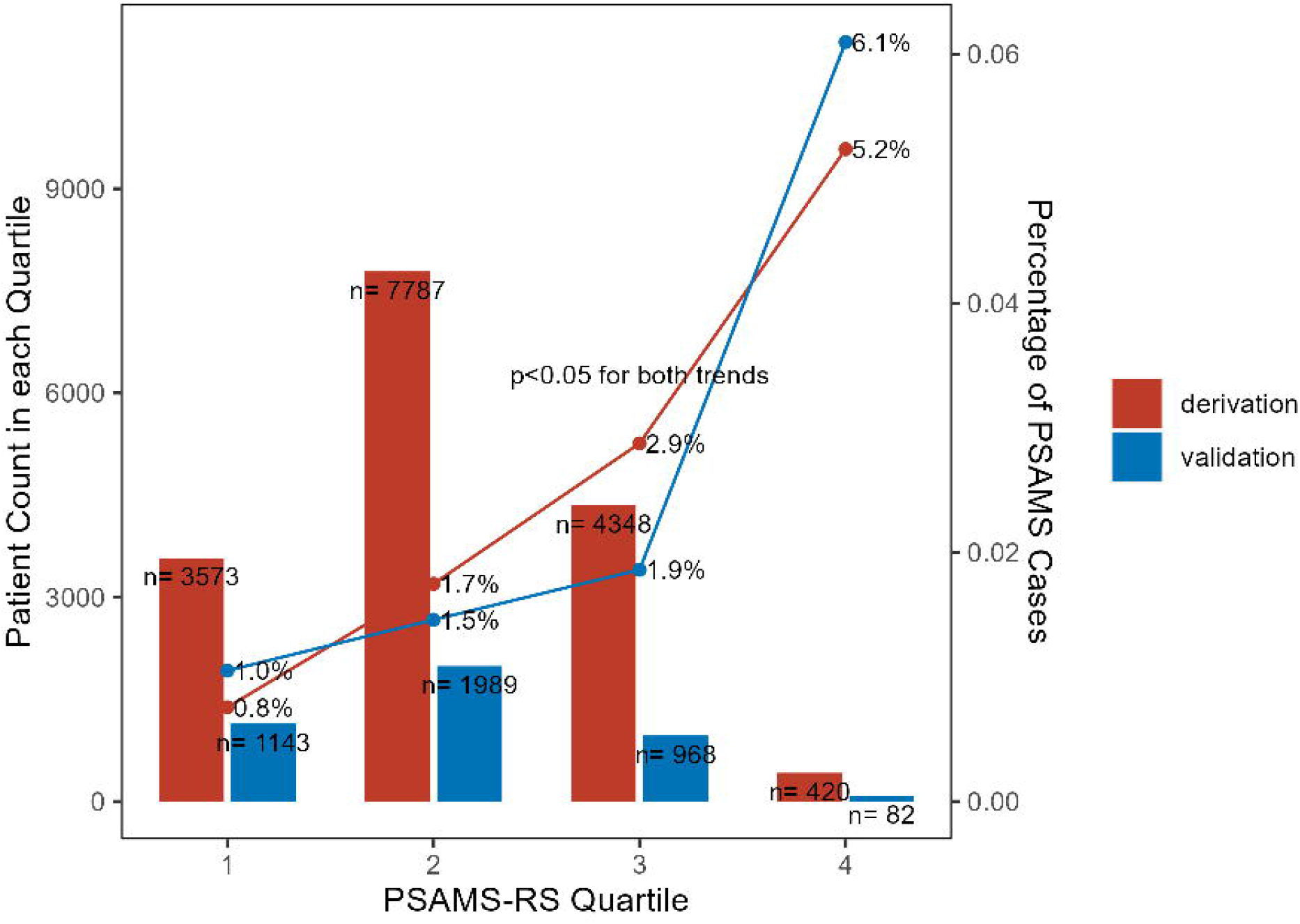
1-Year Pharmacological Statin-Associated Muscle Symptoms (PSAMS) Across Quartile of the PSAMS-RS Score in the Derivation and Validation Cohorts/. Bar plots demonstrate the patient counts in each quartile of PSAMS scores in the derivation and validation cohorts. Trend lines represent the percentage of PSAMS cases in the derivation and validation cohorts across score quartiles.

Figure 2 displays the Kaplan-Meier curves for PSAMS at one year. The PSAMS-RS score effectively stratified patients into higher (score >=102) and lower (score<102) risk groups for PSAMS events in both the derivation (Figure 2B) and validation (Figure 2D) cohorts (log-rank p<0.05 for both). Furthermore, stratifying the risk per PSAMS-RS score quartiles also achieved significant separation in PSAMS risk (log-rank p<0.05 for both) in derivation (Figure 2A) and validation (Figure 2C) cohorts. As shown in Table 3, after adjusting for statin intensity, patients in the 4th quartile for the PSAMS-RS score had an over sevenfold (HR, 7.1; 95% CI, 4.03-12.45) in the derivation and six-fold (HR, 6.1; 95% CI, 2.15-17.45) higher hazard in the validation cohort of developing PSAMS than those in the 1st score quartile.

**Table 3:**
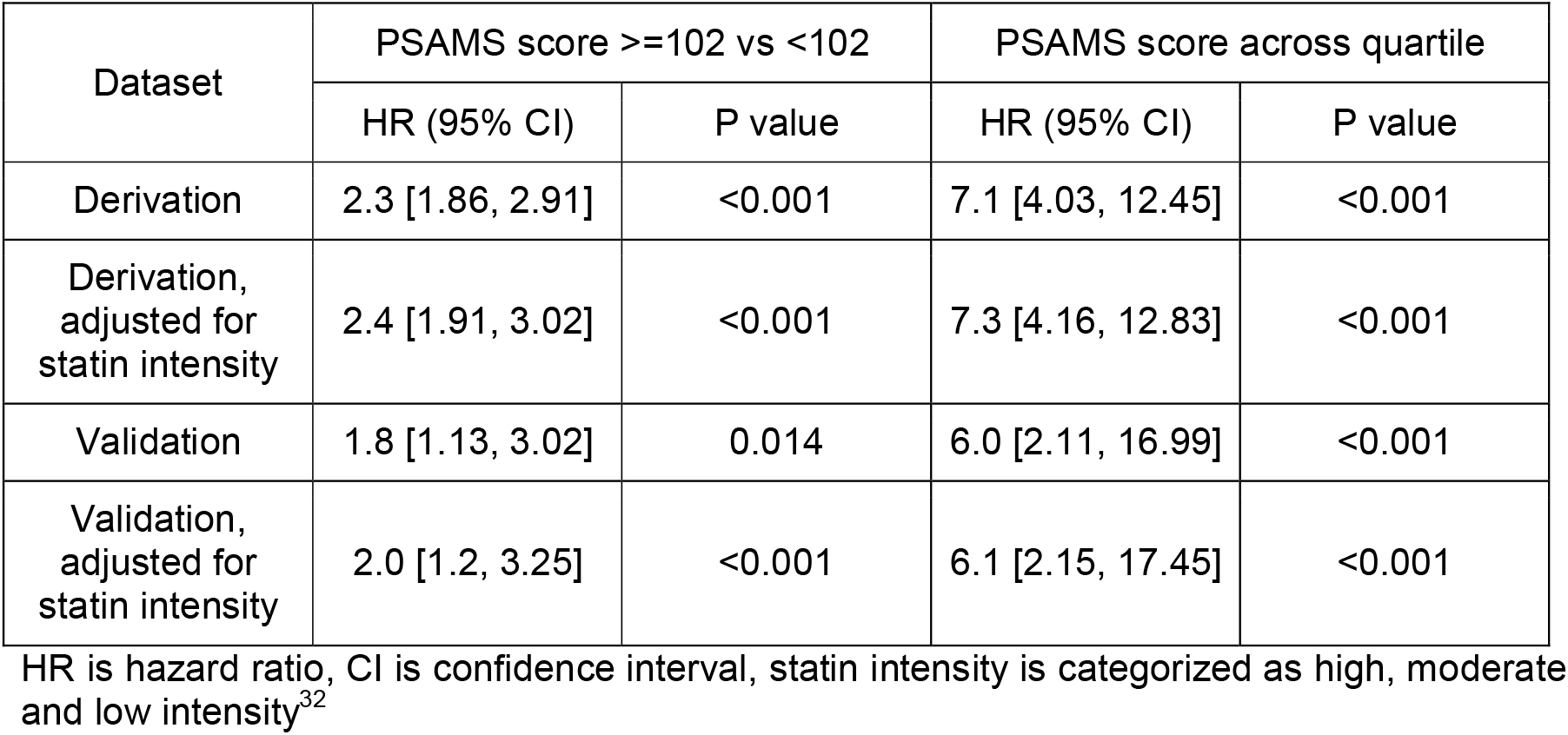
PSAMS Score as a Predictor for 1-year PSAMS Hazard.

**Figure 2:**
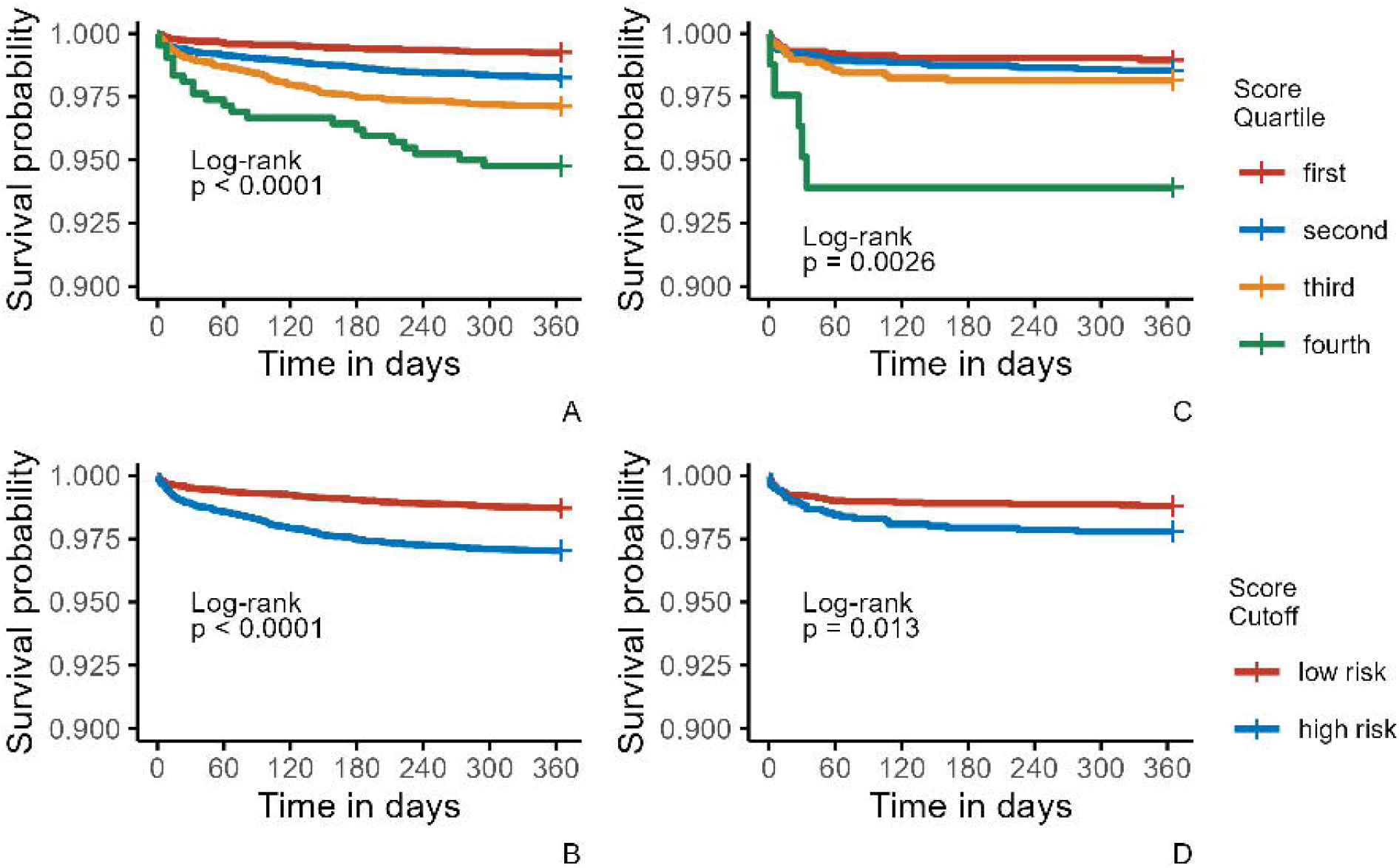
1-Year Pharmacological Statin-Associated Muscle Symptoms (PSAMS) Outcome by PSAMS-RS Score Quartile and Cutoff in Derivation and Validation Cohorts/. Kaplan-Meier curves for PSAMS at 1 year by PSAMS-RS score quartile in the derivation (A) and validation cohorts (C) and; PSAMS score cutoff (<102 (low risk) vs >=102 (high risk)) in the derivation (B) and validation (D) cohorts.

The ROC AUC scores for the PSAMS-RS model was 0.65 (95% CI: 0.62 to 0.68) in the derivation cohort and 0.62 (95% CI: 0.54 to 0.69) in the validation cohort. The Hosmer-Lemeshow test p value was 0.099 (non-significant), indicating good fit of the model. The performance of the PSAMS-RS model score was marginally better than the selected machine learning algorithms as indicated by ROC AUC scores (Figure S1). After applying the MRS^26^ on the derivation set, there was no significant trend of PSAMS prevalence across quartiles (p for trend =0.09) (Figure S2) and no significant separation of Kaplan-Meier curve for PSAMS at one year (log-rank p =0.15) (Figure S3).

## Discussion

Characterizing clinical risk factors associated with pharmacological SAMS (PSAMS) has proven challenging in real-world settings. This has been primarily due to the prevalence of reported SAMS being influenced by nocebo effects.^33,34^ Nevertheless, several common risk factors have been identified to be associated with risk for SAMS, including older age, female sex, Asian ancestry, lower body weight, and the presence of comorbidities and select concurrent medications.^3^

Recent studies have examined the possibility to develop prognostic scores to identify patients with higher risk of SAMS.^26,35^ Hopewell et al. proposed the Myopathy Risk Score (MRS) for simvastatin-associated myopathy based on pooled clinical trial data.^26^ When applying the MRS to our derivation cohort, it did not stratify patients’ risk of developing PSAMS with an useful level of discrimination (Figure S2 and S3), raising concerns of its potentially applicability using data generated from real-world settings. It’s also important to note that the MRS focuses exclusively on simvastatin, while other statins also contribute to PSAMS as well. Another study by Collins et al. introduced the QStatin risk score, but its definition of myopathy outcome primarily relied on diagnosis codes and CK elevations alone, and thus lacking consideration of contributions made by considering unstructured components in EHRs implicating possible PSAMS events.^35^ Additionally, the QStatin score lacked clarity as to the specificity of its predictive power for general myopathy as opposed to specific statin-associated myopathy.

The PSAMS-RS score represents a novel approach to specifically stratify PSAMS risks based on patients’ clinical factors using both structured and unstructured EHR data. The score was developed through the utilization of our validated PSAMS phenotyping algorithm, which specifically identifies PSAMS cases rather than considering overall SAMS including nocebo effects. By focusing on EHR data, we gained the advantage of accessing a substantial volume of real-world data with numerous available clinical variables. The LASSO variable selection process also has proven success in other risk prediction model developments.^22,23^ The findings of the PSAMS-RS score indicate that patients with a score of 102 or higher have a two-fold higher risk of developing PSAMS within one year of initiating statin therapy. Moreover, patients in the 4th quartile of the PSAMS score face an over six-fold higher hazard compared to those in the 1st quartile. This score offers a combinatory and semi-quantitative approach to fine tuning a patient’s risk of PSAMS rather than examining individual risk factors alone.

The PSAMS-RS score has certain limitations. Firstly, it was developed and validated using EHR data collected from 1/1/2010 to 12/31/2018 and 1/1/2019 to 12/31/2020, respectively. Although the temporal validation of prediction models has been adopted in previous studies^17,36,37^, to ensure the generalizability of the score, external validation using data from a different EHR system is necessary. It is also worth noting that the majority of included clinical variables (Table 1) had significantly different distributions between derivation and validation cohorts. Therefore, our successful validation of the PSAMS-RS score in this heterogeneous validation cohort adds confidence in the potential generalizability of the score in external datasets. Secondly, since our focus was specifically on PSAMS, which have a relatively low prevalence in real-world settings (approximately 2%^3^), the dataset used for development was highly unbalanced. Even though the discrimination ability was reasonable (ROC AUC scores were 0.65 in the derivation and 0.62 in the validation cohort), the imbalanced nature of these datasets posed challenges to developing a risk prediction model with high precision (the accuracy of positive predictions for PSAMS cases). Various machine learning algorithms were tried, however the performance measures were slightly worse (Figure S1). Therefore, it is important to interpret the PSAMS-RS score with caution. Rather than solely relying on it as a risk prediction model for estimating the risk of PSAMS, it should be considered as a risk stratification tool that helps in triaging patients based on their risk of PSAMS.

The potential implementation of PSAMS-RS score provides clinicians with a novel clinical informatics tool to better identify patients that potentially could develop PSAMS, thereby enabling the clinician to take preventative measures (e.g. adjustment of target doses, taking drug holidays, reviews of potentially interacting medications and obtaining relevant lab values, etc.) in order to improve statin adherence, and ultimately enhance cardiovascular outcomes such as reductions in morbidity and mortality for patients receiving statins. For future directions, we intend to replicate the PSAMS cohort using another EHR system to validate the generalizability of the score. Additionally, we aim to incorporate pharmacogenomics information, such as SLCO1B1, ABCG2, and CYP2C9 genotypes, into our risk stratification tool. The integration of genetic data is expected to further improve the accuracy of the PSAMS-RS score.^38^

## Conclusion

The PSAMS-RS score offers a straightforward tool for stratifying patients based on their risk of developing PSAMS following statin initiation. Patients at the 4th score quartile had over six-fold higher risk of PSAMS than those in the 1st quartile. The potential utilization of the scores allows clinicians to take proactive measures to prevent potential statin non-adherence associated with PSAMS. External evaluation of the PSAMS-RS score is warranted to further define its generalizability and clinical utility.

## Supporting information

PSAMS-RS Risk Calculator

## Data Availability

All data produced in the present study are available upon reasonable request to the authors

