## Supplemental Material for "Development and validation of the pharmacological statin-associated muscle symptoms risk stratification (PSAMS-RS) score using real-world electronic health record data"

**Table S1:** ICD Codes Definition for the PSAMS-RS Score Comorbidity Variables

| Variable | ICD-CM codes |
| --- | --- |
| Chronic pulmonary disease (Charlson Quan 2005 & Elixhauser Quan 2005) | ICD-9-CM: 490.x-505.x, 416.8x, 416.9x, 506.4x, 508.1x, 508.8x  ICD-10-CM: I27.8x, I27.9x, J40.x-J47.x, J60.x-J67.x, J68.4x, J70.1x, J70.3x |
| Neurological disease (Elixhauser Quan 2005) | ICD-9-CM: 332.x. 334.x-335.x, 340.x-341.x, 345.x, 331.9x, 333.4x-333.5x, 333.92, 336.2x, 348.1x, 348.3x, 780.3x, 784.3x  ICD-10-CM: G10.x-G13.x, G20.x-G22.x, G25.4x-G25.5x, G31.2x, G31.8x-G31.9x, G32.x, G35.x-G37.x, G40.x-G41.x, G93.1x, G93.4x, R47.0x, R56.x |
| Renal disease (Charlson Quan & Elixhauser Quan 2005) | ICD-9-CM: 582.x, 583.0x-583.7x, 585.x, 586.x, 588.x, 403.01, 403.11, 403.91, 404.02, 404.03, 404.12, 404.13, 404.92, 404.93, V42.0x, V45.1x, V56.x  ICD-10-CM: I12.0x, I13.1x, N03.2x-N03.7x, N05.2x-N05.7x, N18.x-N19.x, N25.0x, Z49.0x-Z49.2x, Z94.0x, Z99.2x |
| Cerebrovascular disease (Charlson Quan 2005) | ICD-9-CM: 430.x-438.x, 362.34  ICD-10-CM: G45.x-G46.x, H34.0x, I60.x-I69.x |
| Hypothyroidism (Elixhauser Quan 2005) | ICD-9-CM: 243.x-244.x, 240.9x, 246.1x, 246.8x  ICD-10-CM: E00.x-E03.x, E89.0x |
| Lymphoma (Elixhauser Quan 2005) | ICD-9-CM: 200.x-203.x, 238.6x  ICD-10-CM: C81.x-C85.x, C88.x, C90.0x, C90.2x, C96.x |
| Peripheral vascular disease (Charlson Quan 2005^a,c^ & Elixhauser Quan 2005) | ICD-9-CM: 440.x-441.x, 443.1x-443.9x, 447.1x, 437.3x, 557.1x, 557.9x, 93x, V43.4x  ICD-10-CM: I70.x-I71.x, I73.1x, I73.8x-I73.9x, I77.1x, I79.0x, I79.2x, K55.1x, K55.8x-K55.9x, Z95.8x-Z95.9x |
| Coronary artery disease | ICD-9-CM: 411.x, 414.x (except: 414.10, 414.19)  ICD-10-CM: I24.x, I25.x (except: I25.3) |

ICD is international classification of diseases

**Figure S1: Area Under Receiver Operating Characteristic Curve Comparing Performances of LASSO Regression Versus Other Selective Machine Learning Algorithms in the Testing and Validation Cohorts/** LASSO = Least Absolute Shrinkage and Selection Operator

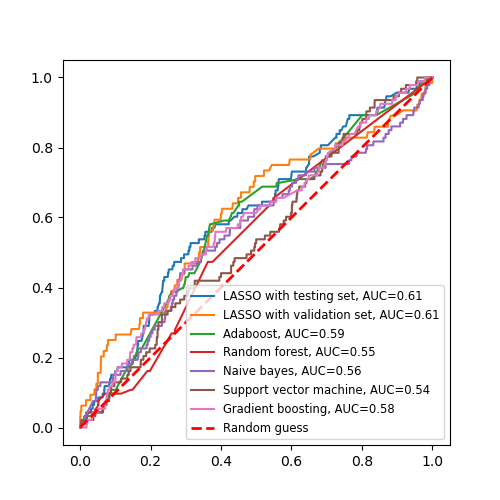

**Figure S2: 1-Year Pharmacological Statin-Associated Muscle Symptoms (PSAMS) Across Quartiles of the Myopathy Risk Score (MRS) in the Derivation Cohort**/ Bar plots demonstrate the patient counts in each quartile of MRS in the derivation cohort. The trend line represents the percentage of PSAMS cases in the derivation cohort across score quartiles.

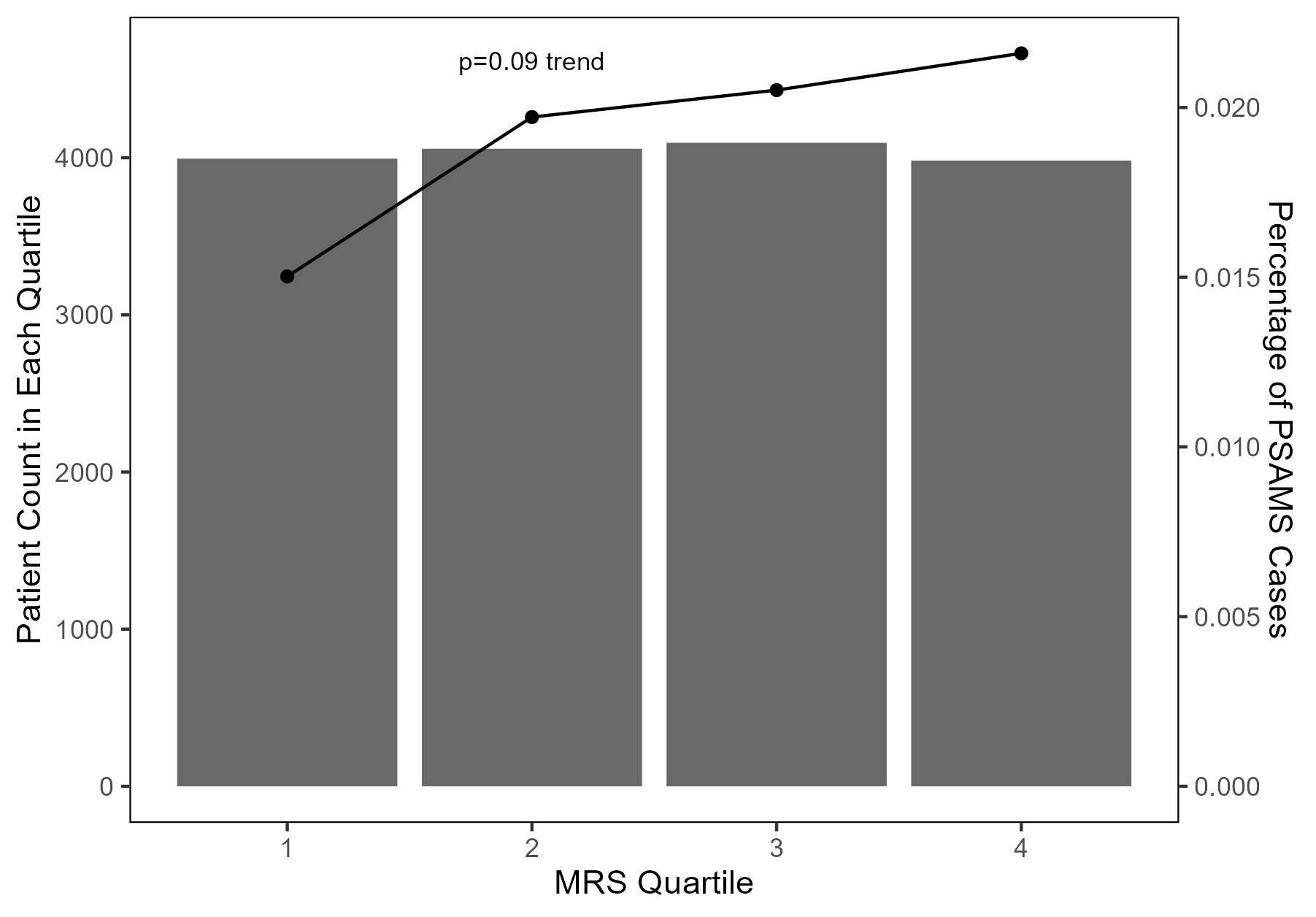

**Figure S3: 1-Year Pharmacological Statin-Associated Muscle Symptoms (PSAMS) Outcome by Myopathy Risk Score (MRS) Quartile in Derivation Cohort**/ The Kaplan-Meier curve for PSAMS at 1 year by MRS quartile in the derivation cohort.

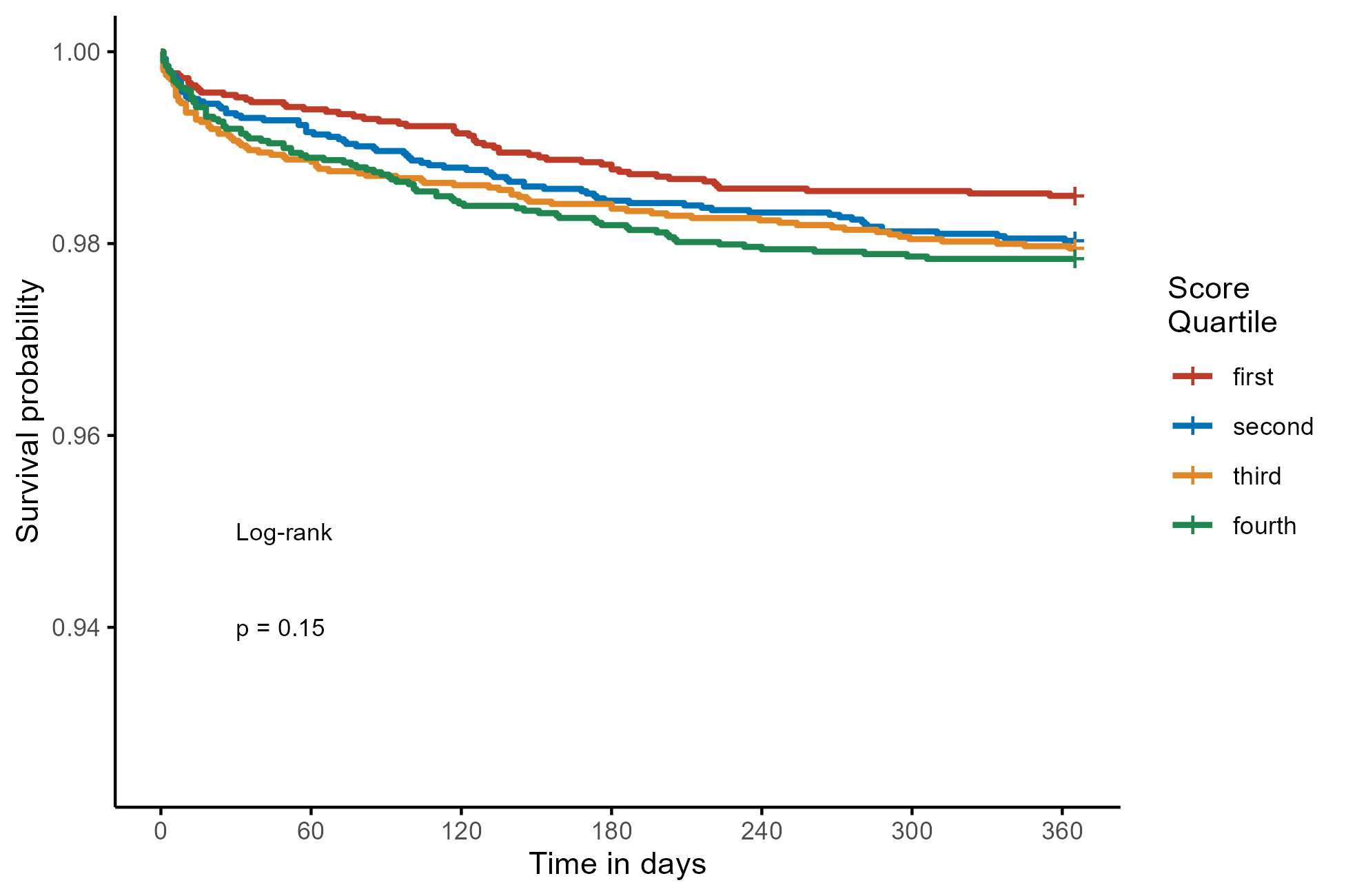
